# The Physical Health Trajectories of Young People with Neurodevelopmental Conditions: A Protocol for a Systematic Review of Longitudinal Studies

**DOI:** 10.1101/2024.03.22.24304709

**Authors:** Naomi Wilson, Ruchika Gajwani, Michael Fleming, Mia Findlay, Helen Stocks, Graham Walker, Naeve Corrigan, Helen Minnis

**Author notes:** Corresponding Author: Dr Naomi Wilson, Address:Clarice Pears Building, 90 Byres Road, University of Glasgow, Glasgow G12 8TB. PROPSPERO Registration Number: CRD42024516684.

## Abstract

**Introduction:** It is now widely acknowledged that without appropriate support, young people with neurodevelopmental conditions (NDCs) are at an increased risk of many of the social and psychiatric outcomes which are known to be key drivers of physical health inequalities. Despite this, until recently relatively little attention has been paid to their physical health trajectories. There is now emerging longitudinal evidence to suggest an association between specific NDCs in childhood or adolescence and certain physical long-term conditions (LTCs) in adulthood. However, to date this literature has never been comprehensively appraised. As a result, our understanding of all the future health risks that young people with NDCs may collectively be at risk of is limited and the factors which drive these adult health outcomes also remain obscure.

**Methods:** A search strategy has been developed in collaboration with two medical librarians and will be used to conduct systematic searches of Medline, Embase, APA PsycINFO, Cumulative Index to Nursing and Allied Health Literature, and Web of Science. Prospective longitudinal studies exploring the association between three common NDCs in childhood or adolescence (i.e., ADHD, Autism, and Tic Disorders <18 years of age) and any physical LTC in adulthood (i.e., > 18 years of age) will be selected through title and abstract review, followed by a full-text review. Data extracted will include definition of exposure and outcome, mediators or moderators investigated, confounders adjusted for, and crude and adjusted effect estimates. Risk of bias assessment will be conducted. Results will be synthesized narratively and if the data allow, a meta-analysis will also be conducted.

**Ethics and dissemination:** Ethics approval is not applicable for this study since no original data will be collected. The results of the review will be widely disseminated locally, nationally, and internationally through peer-reviewed publication, adhering to the PRISMA statement, and conference presentations.

**Article Summary:** *Strengths and Limitations of This Study:*

- To our knowledge, this is the first systematic review synthesising and critically assessing evidence from longitudinal, observational studies on the association between NDCs in childhood or adolescence and physical long-term conditions (LTCs) in adulthood.
- We will conduct a comprehensive search across multiple databases, without publication restrictions and will adhere to Preferred Reporting Items for Systematic Review and Meta-Analysis Protocols (PRISMA-P) recommendations to ensure methodological rigor.
- This study’s focus on prospective longitudinal evidence from observational studies will strengthen the conclusions drawn from results and may facilitate causal inference across studies.
- Depending on its findings, this study may represent a healthier sample of people with NDCs due to studies with significant loss to follow-up.
- We plan to meta-analyse outcome data; however due to possible heterogeneity between studies this may not be appropriate.

## 1. Introduction

The number of young people being diagnosed with a neurodevelopmental condition (NDC), has dramatically increased in recent years [1]. Specifically, there has been a substantial upsurge in the number of children and adolescents being referred to mental health services for suspected Attention Deficit Hyperactivity Disorder (ADHD), Autism and Tic Disorders (i.e., Transient or Chronic Tic Disorders and Tourette’s Syndrome), without an associated intellectual disability [2,3,4,5,6]. While many of these conditions were previously conceptualized as being limited to childhood, they are now known to be lifelong [7]. As such, an accurate understanding of the future health needs of these young people is essential to informing the development and implementation of services and policies that address these.

It is now widely accepted that when left without appropriate support, young people with NDC’s are at an increased risk of many of the social and psychiatric outcomes, which are known to be key drivers of physical health inequalities globally [8,9]. Specifically, recent systematic reviews of longitudinal studies have shown that compared to the general population, young people with NDC’s are more likely to experience a range of mental health difficulties in adulthood, including substance use, anxiety, and depressive disorders [10,11]. Potentially partially due to these comorbidities, alongside a lack of educational support [12], having an NDC in childhood or adolescence can also increase the risk of future unemployment [13], socioeconomic disadvantage [14], and social exclusion [15]. However, despite knowledge of these heightened risks, until recently relatively little research or clinical attention has been paid to the physical health trajectories that young people with NDCs may experience.

### 1.1 Evidence Before this Study

It is well recognised that in childhood, NDCs are associated with a range of physical health conditions and complaints [16]. Specifically, a systematic review of *cross-sectional* studies identified that children with NDC’s are at an increased risk of autoimmune conditions, epilepsy, and gastrointestinal complaints [17]. A growing body of evidence is also now emerging, to demonstrate a *longitudinal* association between specific NDCs in childhood and adolescence and certain physical long-term conditions (LTCs) in adulthood. For example, an increased risk of Type II Diabetes Mellitus has recently been shown amongst adults diagnosed with Tourette’s Syndrome as children [18], while an increased risk of coronary artery disease has been identified amongst those previously diagnosed with ADHD [19]. Epidemiological studies suggest that most people with these common chronic morbidities have at least two other, and frequently more, LTCs [20]. It is also now increasingly understood that NDCs rarely occur in isolation and frequently co-occur [21]. For instance, young people with Autism, are known to have a mean of 3.2 comorbid NDCs [22], and there is significant overlap in the aetiology, symptoms and consequences associated with these conditions [23, 24].

### 1.2 Added Value of the Study

The risk of experiencing *multiple* physical LTCs in adulthood associated with experiencing *any* NDC in childhood and adolescence may therefore be substantial. However, whether this is the case remains unclear, as to date this emerging research area has been hindered by a selective focus on specific childhood NDCs or on particular physical health outcomes. Moreover, the emerging body of longitudinal research which does exist has never been systematically appraised or collated. There would therefore be great value in integrating the results of the existing studies in this area, in order to aid our understanding of all of the future health risks that young people with NDC’s may collectively be at risk of.

In addition, while some of the factors which may account for these associations have been hypothesized, the exact mechanisms which underpin them are unclear and potential targets for early intervention are therefore obscure [25]. For instance, a recent systematic review exploring the association between ADHD and obesity found that while impulsivity, inflammation and sedentary behaviour have all been suggested as mediating factors, results regarding their significance are conflicting [26]. The authors of this review concluded that only through prospective longitudinal studies will we understand the factors driving these associations more accurately. Comprehensively appraising the longitudinal studies which do exist is therefore the critical first step in determining the behavioural, biological, psychosocial, and genetic factors which have so far been implicated, and identify any remaining research or knowledge gaps on underlying mechanisms.

### 1.3 Review Objectives

The purpose of the current review is therefore to systematically collate and appraise the research which has been conducted to date exploring the longitudinal association between three common NDCs (ADHD, Autism, and Tic Disorders) in childhood or adolescence (i.e., <18 years of age) and isolated or multiple physical LTCs in adulthood (i.e., >18 years of age). In doing so, we aim to answer the following research questions.

> (RQ1) Is there existing evidence to suggest a longitudinal association between ADHD, Autism or Tic Disorders in childhood or adolescence and the subsequent development of physical LTCs in adulthood?
>
> (RQ2) Are there any factors (i.e., biological, behavioural, psychosocial, material, structural, or genetic) which have been shown mediate or moderate this association?

For conditions where an appropriate body of homogeneous evidence exists, we will also conduct a meta-analysis to determine what the pooled estimate is, from the existing literature, regarding the risk of each condition or condition category in adulthood among those diagnosed with an included NDC in childhood or adolescence.

## 2. Methods and Analysis

The Preferred Reporting Items for Systematic Review and Meta-Analysis Protocols (PRISMA-P) recommendations have been used to guide the reporting in this systematic review protocol and the Preferred Reporting Items for Systematic reviews and Meta-Analyses (PRISMA) statement (2020) [27] will be used to guide the reporting of the review itself [28]. This systematic review protocol has been registered in the International Prospective Register of Systematic Reviews (PROSPERO) with registration number [CRD42024516684].

### 2.1 Patient and Public Involvement

Patients and the public were involved in the development and scope of this review and will be involved in the synthesis and reporting of its findings. This has been undertaken with the support of a working patient and public involvement and engagement (PPIE) group. We sought PPIE group members thoughts on the reviews objectives and included conditions during an initial focus group discussion. Based on their feedback during further focus groups, we will also ensure that this review reports outcomes of relevance and importance to stakeholders. Patients and members of the public will not be involved in the study selection or extraction phases but where possible, will be involved with the evidence synthesis stage. We will also seek the guidance of PPIE group members prior to disseminating our findings, via a lay summary, peer reviewed publication and conference presentations, to ensure translatability of results.

### 2.2 Eligibility Criteria

Study selection and eligibility criteria will be based on the Population, Exposure, Comparison, Outcome, Study design (PICOS) outlined below (Table 1).

**Table 1.**
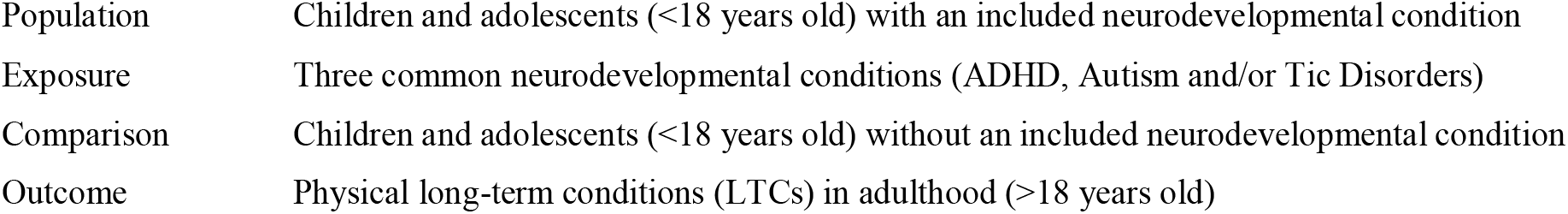
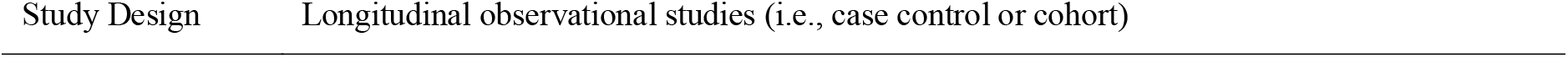
PECO Criteria.

We will search for observational, prospective longitudinal studies, from any country, which have explored the prevalence of isolated physical LTCs and/or multimorbidity among adults diagnosed with an included NDC in childhood or adolescence. Studies will subsequently be deemed eligible for inclusion if they adhere to the inclusion and exclusion criteria outlined below.

#### 2.2.1 Inclusion Criteria

1. Prospective longitudinal studies exploring the association between an *incident* included NDC identified in childhood or adolescence (i.e., before the age of 18) and any *subsequent* physical LTCs present in adulthood (i.e., over the age of 18).
2. To minimise heterogeneity within our sample population, studies will be required to have adopted a systematic approach to the identification of NCD symptoms as defined by the relevant DSM or ICD criteria, or through use of a research diagnostic protocol that matched these standards by considering symptom range, threshold, presence of impairment, chronicity, and pervasiveness.
3. To ensure studies can be considered as beginning in childhood or adolescence only those where participants had a mean age of less than 18 years at baseline will be included.
4. To ensure studies are longitudinal in nature and that participants could reasonably be considered adults at follow-up, only studies where a proportion of participants were greater than 18 years at follow-up will be included.
5. The outcome conditions of interest are any physical LTCs occurring in adulthood (i.e., over the age of 18). As per the National Institute of Health Research (NIHR) recommendations, this will be defined as any health problem that requires ongoing management over a period of years and which cannot currently be cured but can be controlled with the use of medication and/or other therapies.
6. Studies which include any cancer as an outcome will also be included, as this is now widely considered to be a LTC.
7. Publication in English in a book or peer-reviewed journal

#### 2.2.2 Exclusion Criteria

1. Cross-sectional studies.
2. Only studies which include a comparison sample of individuals without an NDC identified in childhood or adolescence, either through inclusion of a reference population or a control group will be included. Studies which do not include a comparison to a non-exposed control group (i.e., case series) will be excluded.
3. Drug trials which primarily focus on the outcomes associated with a single medication for an included NDC will be excluded.
4. As the interest of the current review is on the development of LTCs across the life course, studies which focus on incident or comorbid congenital conditions (i.e., congenital cardiac malformations, cerebral palsy, phenylketonuria, etc.) will also be excluded.
5. Any studies who only report on physical outcomes which are not considered to meet the above definition of an LTC (i.e., such as acute infections, accidents, or injuries) will be excluded.
6. As our focus is on the physical health trajectories associated with our exposure of interest, studies which only report on psychiatric or mental health outcomes will also be excluded.

### 2.3 Information Sources

Databases searched electronically will be MEDLINE, PsycINFO, EMBASE, CINHAL Plus, and Web of Science (WoS). We have selected MEDLINE, EMBASE and WoS databases based on recommendations for Cochrane Handbook covering major health sciences topics. We also selected specialist subject databases CINAHL Plus and PsycINFO based on their relevance to the study objectives. WHO’s trials portal and clinicaltrials.gov will be searched to identify unpublished studies or studies still ongoing. We shall conduct a manual search of the reference lists and forward and backward citation of all included studies using the Web of Science database for any additional papers. We shall exclude studies that are published systematic or literature reviews but shall also examine the reference lists of relevant reviews for additional candidate studies.

### 2.4 Search Strategy

The final search strategy was developed in collaboration with two medical librarians and is included in Appendix 1.

Our exposure of interest was common NDCs and we therefore chose for ADHD, Autism and/or Tic Disorders to be the focus of this review. Search terms for the NDCs of interest were drawn from recent Cochrane reviews for these conditions [29, 30, 31].

Our outcome of interest are any physical LTCs occurring in adulthood. Therefore, we used broad search terms for physical LTCs generally. However, in order for our search strategy to be as comprehensive as possible, we also included search terms for the four conditions which account for the greatest number of DALY’s due to physical LTCs in High-Income Countries where the majority of NDC studies have been conducted to date (i.e., cardiovascular disease, stroke, diabetes, painful musculoskeletal conditions and chronic obstructive pulmonary disease) [32]. In addition, we included search terms for any other conditions which have been repeatedly identified as the most prevalent co-occurring physical LTCs among adults with an incident common mental health condition generally (hypertension, hyperlipidaemia, obesity, peptic ulcer disease, irritable bowel syndrome and chronic pain) [33,34,35,36,37] or among those with Autism, ADHD or a Tic Disorder specifically (asthma, atopic dermatitis, allergic rhinitis, acne, enthesopathy, migraine and epilepsy) [38].

Broad search terms for physical LTC’s generally and for the majority of included LTCs were drawn from the most recent Cochrane review of multimorbidity [39]. Specific search terms for the remaining LTCs were drawn from previously published reviews on chronic pain [40,41], epilepsy [42], obesity [43], allergic rhinitis [44], peptic ulcer disease [45], irritable bowel syndrome [46], atopic dermatitis [47], and acne [48]. Search terms for longitudinal studies were drawn from the British Medical Journal Best Practice Guidelines [49]. These key terms will be combined with Boolean logic to search for relevant studies and medical subject headings will be used to capture concepts and maximise the number of studies retrieved.

### 2.5 Study Records

#### 2.5.1 Data Management

Titles and abstracts of studies retrieved from each database search will be exported to EndNote (version X9) for deduplication, and then imported to Rayyan software for screening.

#### 2.5.2 Study Selection Process

Two review authors will independently screen the titles and abstracts of all identified studies to identify those that meet the predefined inclusion/exclusion criteria as outlined above. The full text of all remaining potentially eligible studies will be independently screened by two authors thereafter. Full texts will also be obtained where necessary to further screen for eligibility if information in the studies title and abstract is insufficient to determine this, or in case of disagreement. Where consensus on eligibility cannot be achieved, a third review author will be involved in the discussion.

#### 2.5.3 Data Extraction

Using a standardised data collection form, two reviewers will independently extract data from the eligible studies. For included studies this will include the author, year of publication, country of origin, study design, sample size, mean age of sample at study start and end, and/or mean period of follow-up, definition of exposure (i.e. NDC assessment method) and outcome (i.e. physical LTC assessment method), mediators or moderators investigated, confounders adjusted for (if any), crude and adjusted effect estimates (with 95% CI) and estimates of subgroup analysis (if any). Discrepancies will be resolved by a third reviewer if necessary.

### 2.6 Outcomes and Prioritization

Our primary outcome of interest will be the risk of physical LTCs in adulthood among those diagnosed with an NDC in childhood and this will be prioritized. Our secondary outcome of interest are any factors which have been identified to mediate or moderate this association. Data on any mediators or moderators involved in this association will subsequently be extracted from studies which also include this and will be analysed separately.

### 2.7 Risk of Bias Assessment

Quality assessment of the included studies will be conducted by two reviewers independently using the Newcastle-Ottawa Scale (NOS) [50]. The NOS is a widely used measure designed to assess the quality of observational studies, with versions available for both case–control and cohort studies. It is currently a tool recommended by the Cochrane Collaboration, and assesses studies according to eight items, categorised into three dimensions: selection, comparability, and outcome or exposure for cohort studies or case–control studies respectively. Studies can be graded up to one point for all items, except comparability which has the potential to score up to two points, giving a maximum possible score of nine. Studies will be rated from 0–9 according to this scale by both reviewers and scores will subsequently be compared and discussed. Any discrepancies will be resolved by a third reviewer if necessary. The agreed upon total scores will then be used to indicate the overall quality of each included study, with those studies rating 0–2 being identified as of poor quality, 3–5 as of medium quality, and 6–9 as of high quality.

#### 2.8.1 Data Synthesis

We will tabulate key information (e.g., sample characteristics, exposure, and outcome characteristics) regarding the association between included NDCs and physical LTCs grouped by the body systems outlined in Table 2 (i.e., cardiovascular, metabolic and endocrine, respiratory, gastrointestinal, musculoskeletal and dermatological) and perform a narrative synthesis regarding these associations. In this narrative synthesis, we will attempt to explore differences and similarities in outcomes between the included NDCs and will also explore the possibility of reporting results for different socio-demographic (e.g., by ethnicity) or other potentially high-risk sub-groups (i.e., looked after or accommodated children).

**Table 2.** Adult LTCs included in search terms.

| Body system (based on ICD-10) | Conditions Included |
| --- | --- |
| Neurological | Chronic Pain, Migraine, Epilepsy |
| Cardiovascular | Cardiovascular Disease, Cardiac Arrhythmias, Hypertension, Stroke |
| Metabolic and Endocrine | Hyperlipidaemia, Obesity, Diabetes Mellitus |
| Respiratory | Asthma, Allergic Rhinitis, Chronic Obstructive Pulmonary Disease |
| Gastrointestinal | Peptic Ulcer Disease/Dyspepsia, Irritable Bowel Syndrome |
| Musculoskeletal | Enthesopathy |
| Dermatological | Atopic Dermatitis, Acne |

We will perform a separate narrative synthesis of any identified mediating or moderating factors in this association and present these results separately. Specifically, we will categorise any factors which have been found to influences this association according to either one of the recognised pathways to inequalities in health [51], or to any biological or genetic pathways identified through our search (Table 3).

**Table 3.** Potential Causal Mechanisms.

| Potential Pathway | Potential Mediating Factors |
| --- | --- |
| Behavioural | High risk health behaviours, including dietary habits, alcohol or illicit substance use and tobacco smoking. |
| Psychosocial | Adverse childhood experiences, or comorbid mental health difficulties. |
| Material | Educational attainment, or socioeconomic status. |
| Structural | Access to health services and resources. |
| Biological | Neural Pathways, Autonomic Nervous System markers, or Inflammatory markers (C-Reactive Protein, Tumour Necrosis Factor alpha) |
| Genetic | Predisposing genetic factors (i.e., SNPs), or epigenetic alterations. |

Where possible and appropriate, meta-analyses will also be performed to calculate the overall pooled estimates of the relationship between the included NDCs combined, and different physical LTCs in adulthood. Both crude and adjusted results will be displayed where possible using the generic inverse variance method. Adjustment will be based on the definition outlined in each identified study and a hierarchy of adjustment created depending on the factors that are adjusted for. Studies that report similar adjustments will be analysed separately in crude and adjusted models to assess potential confounding among studies that reported adjusted estimates.

A fixed-effects model will be used where heterogeneity is low (I2 value of less than 50%), and a random-effects model where heterogeneity is high (I2 value of 50% or more) according to the Cochrane Handbook criteria [52].

#### 2.8.2 Additional Analyses

If appropriate a funnel plot will be used to explore the existence of publication bias by visual inspection. We will also undertake an objective assessment of publication bias using Egger’s regression test. Asymmetry of the funnel plot and/or statistical significance of Egger’s regression test (p<0.05) will be indicative of publication bias.

### 2.9 Confidence in cumulative evidence

At least two independent reviewers will assess the strength of the body of evidence (according to the following categories: high, moderate, low and very low) for each individual outcome using the five Grades of Recommendations, Assessment, Development and Evaluation (GRADE) considerations for downgrading the certainty of evidence (risk of bias, impression, inconsistency, indirectness and publication bias) and the three GRADE considerations for upgrading the certainty of evidence (large magnitude of effect, dose-response gradient and residual confounding decreasing the overall effect) [53].

## 3. Ethics and Dissemination

Ethics is not required for this study, given that this is a protocol for a systematic review, which uses published data. The results of the review will be widely disseminated locally, nationally, and internationally. A paper will be submitted to a leading peer-reviewed journal in this field, and reporting of the study will adhere to the PRISMA statement. The findings shall also be presented at a relevant international conference.

## Author Contributions

- Dr Naomi Wilson: study concept and design, data collection, analysis and interpretation, writing and funding acquisition.
- Dr Ruchika Gajwani: study concept and design, interpretation, and supervision
- Dr Michael Fleming: study concept and design, interpretation, and supervision
- Mia Findlay: data collection, analysis, assembly and interpretation
- Helen Stocks: data collection, analysis, and assembly
- Dr Graham Walker: data analysis
- Dr Naeve Corrigan: data analysis
- Professor Helen Minnis: study concept and design, interpretation, and supervision

## Data Availability Statement

No data was collected in the completion of this work.

## Appendix 1. Search Terms

|  | MEDLINE (Ovid) | EMBASE (Ovid) | PSYCINFO (EBSCO) | CINHAL (EBSCO) | Web Of Science (Core Collection) |
| --- | --- | --- | --- | --- | --- |
| **Search Category 1. Neurodevelopmental Disorder Search Terms** |  |  |  |  |  |
| 1 | exp Attention Deficit Disorder with Hyperactivity/ | exp attention deficit disorder/ or exp attention deficit hyperactivity disorder/ | exp Attention Deficit Disorder with Hyperactivity/ | MH "Attention Deficit Hyperactivity Disorder" | (TI=( "Attention Deficit Hyperactivity Disorder" )) OR AB=( "Attention Deficit Hyperactivity Disorder" ) |
| 2 | (ADHD or ADDH or ADHS or "AD/HD" or HKD or TDAH).ti,ab. | (ADHD or ADDH or ADHS or "AD/HD" or HKD or TDAH).ti,ab. | (ADHD or ADDH or ADHS or "AD/HD" or HKD or TDAH).ti,ab. | TI ( (ADHD or ADDH or ADHS or "AD/HD" or HKD or TDAH) ) or AB ( (ADHD or ADDH or ADHS or "AD/HD" or HKD or TDAH) ) | TI=((ADHD or ADDH or ADHS or "AD/HD" or HKD or TDAH) ) OR AB=((ADHD or ADDH or ADHS or "AD/HD" or HKD or TDAH) ) |
| 3 | ((attention* or behav*) adj3 (defic* or dysfunc* or disorder*)).ti,ab. | ((attention* or behav*) adj3 (defic* or dysfunc* or disorder*)).ti,ab. | ((attention* or behav*) adj3 (defic* or dysfunc* or disorder*)).ti,ab. | TI ((attention* or behav*) n2 (defic* or dysfunc* or disorder*)) or AB ((attention* or behav*) n2 (defic* or dysfunc* or disorder*)) | TI=((((attention* or behav*) NEAR/2 (defic* or dysfunc* or disorder*)) ) ) OR AB=((((attention* or behav*) NEAR/2 (defic* or dysfunc* or disorder*)) ) ) |
| 4 | ((disrupt* adj3 disorder*) or (disrupt* adj3 behav*) or (defian* adj3 disorder*) or (defian* adj3 behav*)).ti,ab. | ((disrupt* adj3 disorder*) or (disrupt* adj3 behav*) or (defian* adj3 disorder*) or (defian* adj3 behav*)).ti,ab. | ((disrupt* adj3 disorder*) or (disrupt* adj3 behav*) or (defian* adj3 disorder*) or (defian* adj3 behav*)).ab,ti. | TI ((disrupt* n2 disorder*) or (disrupt* n2 behav*) or (defian* n2 disorder*) or (defian* n2 behav*)) or AB ((disrupt* n2 disorder*) or (disrupt* n2 behav*) or (defian* n2 disorder*) or (defian* n2 behav*)) | AB ((disrupt* n2 disorder*) or (disrupt* n2 behav*) or (defian* n2 disorder*) or (defian* n2 behav*)) or TI ((disrupt* n2 disorder*) or (disrupt* n2 behav*) or (defian* n2 disorder*) or (defian* n2 behav*)) |
| 5 | (impulsiv* or inattentiv* or inattention*).ti,ab. | (impulsiv* or inattentiv* or inattention*).ti,ab. | (impulsiv* or inattentiv* or inattention*).ab,ti. | TI (impulsiv* or inattentiv* or inattention*) or AB (impulsiv* or inattentiv* or inattention*) | TI=((impulsiv* or inattentiv* or inattention*) ) ) or AB=((impulsiv* or inattentiv* or inattention*) ) |
| 6 | (hyperkin* or hyper adj kin*).ti,ab. | (hyperkin* or hyper adj kin*).ti,ab. | (hyperkin* or hyper adj kin*).ti,ab. | TI (hyperkin* or hyper n1 kin*) or AB (hyperkin* or hyper n1 kin*) | TI=(( hyperkin* or hyper NEAR/1 kin*) ) ) or AB=(( hyperkin* or hyper NEAR/1 kin*) ) |
| 7 | (hyperactiv* or hyper adj activ*).ti,ab. | (hyperactiv* or hyper adj activ*).ti,ab. | (hyperactiv* or hyper adj activ*).ti,ab. | TI (hyperactiv* or hyper n1 activ*) or AB (hyperactiv* or hyper n1 activ*) | TI=(( hyperactiv* or hyper NEAR/1 activ*) ) ) or AB=(( hyperactiv* or hyper NEAR/1 activ*) ) |
| 8 | exp Hyperkinesia/ | exp hyperkinesia/ | exp Hyperactivity/ | (MH "Hyperkinesia") | (TI=Hyperkinesia or AB=Hyperkinesia) |
| 9 | (minimal adj brain adj3 disorder*).ti,ab. | (minimal adj brain adj3 disorder*).ti,ab. | (minimal adj brain adj3 disorder*).ab,ti. | TI (minimal n2 brain n2 (disorder*)) or AB (minimal n2 brain n2 (disorder*)) | TI=(minimal NEAR/1 brain NEAR/1 (disorder*)) or AB=(minimal NEAR/1 brain NEAR/1 (disorder*)) |
| 10 | (minimal adj brain adj3 dysfunction*).ti,ab. | (minimal adj brain adj3 dysfunction*).ti,ab. | (minimal adj brain adj3 dysfunction*).ab,ti. | TI (minimal n2 brain n2 (dysfunc*)) or AB (minimal n2 brain n2 (dysfunc*)) | TI=(minimal NEAR/1 brain NEAR/1 (dysfunc*)) or AB=(minimal NEAR/1 brain NEAR/1 (dysfunc*)) |
| 11 | (minimal adj brain adj3 damage*).ti,ab. | (minimal adj brain adj3 damage*).ti,ab. | (minimal adj brain adj3 damage*).ab,ti. | TI (minimal n2 brain n2 (damage*)) or AB (minimal n2 brain n2 (damage*)) | TI=(minimal NEAR/1 brain NEAR/1 (damage*)) or AB=(minimal NEAR/1 brain NEAR/1 (damage*)) |
| 12 | exp Child Development Disorders, Pervasive/ or exp Autistic Disorder/ | exp autism/ | exp Autism Spectrum Disorders/ | MH "Child Development Disorders, Pervasive+" | (TI=( "Child Development Disorders" )) OR AB=( "Child Development Disorders" ) |
| 13 | pervasive development* disorder*.ti,ab. | pervasive development* disorder*.ti,ab. | pervasive development* disorder*.ti,ab. | AB ( "pervasive development* disorder*" ) or TI ( "pervasive development* disorder*" ) | (TI=(( "pervasive development* disorder*" ) ) ) OR AB=(( "pervasive development* disorder*" ) ) |
| 14 | (PDD or PDDs or ASD or ASDs).ti,ab. | (PDD or PDDs or ASD or ASDs).ti,ab. | (PDD or PDDs or ASD or ASDs).ti,ab. | AB (PDD or PDDs or ASD or ASDs) or TI (PDD or PDDs or ASD or ASDs) | (TI=((PDD or PDDs or ASD or ASDs) ) ) OR AB=((PDD or PDDs or ASD or ASDs) ) |
| 15 | autis*.ti,ab. | autis*.ti,ab. | autis*.ti,ab. | TI (autis*) or AB (autis*) | (TI=(autism)) OR AB=(autism) |
| 16 | asperger*.ti,ab. | asperger*.ti,ab. | asperger*.ti,ab. | TI (asperger*) or AB (asperger*) | (TI=((asperger*) ) ) OR AB=((asperger*) ) |
| 17 | kanner*.ti,ab. | kanner*.ti,ab. | kanner*.ti,ab. | TI (kanner*) or AB (kanner*) | (TI=(kanner*)) OR AB=(kanner*) |
| 18 | childhood schizophreni*.ti,ab. | childhood schizophreni*.ti,ab. | childhood schizophreni*.ti,ab. | TI ( "childhood schizophren*" ) or AB ( "childhood schizophren*" ) | AB ( "childhood schizophren*" ) or TI ( "childhood schizophren*" ) |
| 19 |  |  |  |  |  |
| 20 | exp Tourette Syndrome/ or Tourette*.ti,ab. | exp Gilles de la Tourette syndrome/ or Tourette*.ti,ab. | exp Tourette Syndrome/ or Tourette*.ti,ab. | MH "Tourette Syndrome" or TI (Tourette*) or AB (Tourette*) | TI=(Tourette*) or AB=(Tourette*) |
| 21 | exp Tics/ or exp Tic Disorders/ or (tic or tics).ti,ab. | exp tic/ or (tic or tics).ti,ab. | exp Tics/ or exp Tic Disorders/ or (tic or tics).ti,ab. | MH "Tic" or TI (tic or tics) or AB (tic or tics) | AB (tic*) or TI (tic*) |
| **Search Category 2. Physical LTC Search Terms** |  |  |  |  |  |
| 22 | exp Chronic Disease/ | exp chronic disease/ | exp Chronic Illness/ | (MH "Chronic Disease+") | (TI=( chronic* NEAR/3 disease*)) OR AB=( chronic* NEAR/3 disease*) |
| 23 | ((chronic* or "long term") adj3 (illness or disease* or disorder* or condition*)).ti,ab. | ((chronic* or "long term") adj3 (illness or disease* or disorder* or condition*)).ti,ab. | ((chronic* or "long term") adj3 (illness or disease* or disorder* or condition*)).ti,ab. | TI ((chronic* or "long term" or "long-term") n3 (illness or disease* or disorder* or condition*)) or AB ((chronic* or "long term" or "long-term") n3 (illness or disease* or disorder* or condition*)) | TI=((("long term" or "long-term") NEAR/3 (illness or disease* or disorder* or condition*)) ) ) OR AB=((("long term" or "long-term") NEAR/3 (illness or disease* or disorder* or condition*)) ) |
| 24 | exp Epilepsy/ | exp Epilepsy/ | exp Epilepsy/ | MH "Epilepsy" | (TI=(epilep*) ) OR AB=(epilep*) |
| 25 | exp Seizures/ | exp Seizures/ | exp Seizures/ | MH "Seizures" | (TI=(seizure*)) OR AB=(seizure*) |
| 26 | (epilep* or seizure* or convuls*).ti,ab. | (epilep* or seizure* or convuls*).ti,ab. | (epilep* or seizure* or convuls*).ti,ab. | TI (epilep* or seizure* or convuls*) or AB (epilep* or seizure* or convuls*) | (TI=(convuls*)) OR AB=(convuls*) |
| 27 | exp pain/ | exp pain/ | exp pain/ | MH "pain" | - |
| 28 | (chronic adj2 pain*).ti,ab. | (chronic adj2 pain*).ti,ab. | (chronic adj2 pain*).ti,ab. | TI (chronic n2 pain*) or AB (chronic n2 pain*) | AB (chronic NEAR/2 pain*) or TI (chronic NEAR/2 pain*) |
| 29 | (chronic adj2 (discomfort or ache* or neuralgi*).ti,ab. | (chronic adj2 (discomfort or ache* or neuralgi*).ti,ab. | (chronic adj2 (discomfort or ache* or neuralgi*).ti,ab. | TI (chronic n2 (discomfort or ache* or neuralgi*)) or AB (chronic n2 (discomfort or ache* or neuralgi*)) | AB (chronic NEAR/2 (discomfort or ache* or neuralgi*)) or TI (chronic NEAR/2 (discomfort or ache* or neuralgi*)) |
|  | exp headache/ or exp migraine/ | exp headache/ or exp migraine/ | exp headache/ or exp migraine/ |  | - |
| 30 | (headache* or migraine* or neuralgia*).ti,ab. | (headache* or migraine* or neuralgia*).ti,ab. | (headache* or migraine* or neuralgia*).ti,ab. | TI (headache* or migraine* or neuralgia*) or AB (headache* or migraine* or neuralgia*) | (TI=((headache* or migraine* or neuralgia*))) OR AB=((headache* or migraine* or neuralgia*)) |
| 31 | exp heart diseases/ or exp cardiovascular disease/ | exp heart disease/ or exp cardiovascular disease/ | exp Heart Disorders/ | MH "Heart Diseases+" or MH "cardiovascular disease" | - |
| 32 | ((heart or cardiac or cardiovascular or coronary) adj (disease* or disorder* or failure)) or arrythmia*).ti,ab. | ((heart or cardiac or cardiovascular or coronary) adj (disease* or disorder* or failure)) or arrythmia*).ti,ab. | ((heart or cardiac or cardiovascular or coronary) adj (disease* or disorder* or failure)) or arrythmia*).ti,ab. | TI (((heart or cardiac or cardiovascular or coronary) n1 (disease* or disorder* or failure)) or arrythmia*) or AB ((heart or cardiac or cardiovascular or coronary) n1 (disease* or disorder* or failure)) or arrythmia*) | (TI=((((heart or cardiac or cardiovascular or coronary) NEAR/1 (disease* or disorder* or failure)) or arrythmia*))) OR AB=((((heart or cardiac or cardiovascular or coronary) NEAR/1 (disease* or disorder* or failure)) or arrythmia*)) |
| 33 | exp hypertension/ | exp hypertension/ | exp hypertension/ | MH Hypertension | - |
| 34 | (hypertens* or "high blood pressure").ti,ab. | (hypertens* or "high blood pressure").ti,ab. | (hypertens* or "high blood pressure").ti,ab. | TI (hypertens* or "high blood pressure") or AB (hypertens* or "high blood pressure") | (TI=((hypertens* or "high blood pressure*"))) OR AB=((hypertens* or "high blood pressure*")) |
| 35 | exp stroke/ | exp cerebrovascular accident/ or exp stroke/ | exp Cerebrovascular Accidents/ | MH stroke | - |
| 36 | (stroke or strokes or cva or "cerebrovascular accident").ti,ab. | (stroke or strokes or cva or "cerebrovascular accident").ti,ab. | (stroke or strokes or cva or "cerebrovascular accident").ti,ab. | TI ((stroke or strokes or cva or "cerebrovascular accident") or AB ((stroke or strokes or cva or "cerebrovascular accident")) | (TI=((((stroke or strokes or cva or "cerebrovascular accident*"))) OR AB=((stroke or strokes or cva or "cerebrovascular accident*")) |
| 37 | exp diabetes mellitus/ | exp diabetes mellitus/ | exp diabetes mellitus/ | MH diabetes mellitus | - |
| 38 | diabet*.ti,ab. | diabet*.ti,ab. | diabet*.ti,ab. | TI (diabet*) or AB (diabet*) | (TI=(diabet*)) OR AB=(diabet*) |
| 39 | exp hyperlipidemia/ | exp hyperlipidemia/ | exp lipid metabolism disorders/ | MH hyperlipidemia | - |
| 40 | (hyperlipidem* or hypercholesterolemia* or hypertriglyceridemia*).ti,ab. | (hyperlipidem* or hypercholesterolemia* or hypertriglyceridemia*).ti,ab. | (hyperlipidem* or hypercholesterolemia* or hypertriglyceridemia*).ti,ab. | TI (hyperlipidem* or hypercholesterolemia* or hypertriglyceridemia*) or AB (hyperlipidem* or hypercholesterolemia* or hypertriglyceridemia*) | (TI=((hyperlipidem* or hypercholesterolemia* or hypertriglyceridemia*))) OR AB=((hyperlipidem* or hypercholesterolemia* or hypertriglyceridemia*)) |
| 41 | exp Obesity/ or exp Obesity, Morbid/ | exp obesity/ or exp morbid obesity/ | exp Obesity/ | (MH "Obesity+" ) OR (MH "Obesity, Morbid") | - |
| 42 | (adipos* or obes*).ti,ab. | (adipos* or obes*).ti,ab. | (adipos* or obes*).ti,ab. | TI (adipos* or obes*) or AB (adipos* or obes*) | (TI=((adipos* or obes*))) OR AB=((adipos* or obes*)) |
| 43 | exp asthma/ | exp asthma/ | exp Asthma/ | (MH "Asthma+" ) | - |
| 44 | asthma*.ti,ab. | asthma*.ti,ab. | asthma*.ti,ab. | TI (asthma*) or AB (asthma*) | (TI=(asthma*)) OR AB=(asthma*)) |
| 45 | exp pulmonary disease chronic obstructive/ | exp chronic obstructive lung disease/ | exp Chronic Obstructive Pulmonary Disease/ | (MH "Pulmonary Disease, Chronic Obstructive") | (TI=("Chronic Obstructive Pulmonary Disease")) OR AB=("Chronic Obstructive Pulmonary Disease")) |
| 46 | (copd or (pulmonary adj2 (disease* or disorder*))).ti,ab. | (copd or (pulmonary adj2 (disease* or disorder*))).ti,ab. | (copd or (pulmonary adj2 (disease* or disorder*))).ti,ab. | TI (copd or (pulmonary n2 (disease* or disorder*))) or AB (copd or (pulmonary n2 (disease* or disorder*))) | (TI=((copd or (pulmonary NEAR/2 (disease* or disorder*)))) OR AB=((copd or (pulmonary NEAR/2 (disease* or disorder*)))) |
| 47 | exp allergic rhinitis/ or exp rhinitis/ | exp allergic rhinitis/ or exp rhinitis/ | exp Allergic Disorders/ | MH "Rhinitis" | - |
| 48 | rhinit*.ti,ab. | Rhinit*.ti,ab. | Rhinit*.ti,ab. | TI (Rhinit*) or AB (Rhinit*) | (TI=(Rhinit*)) OR AB=(Rhinit*) |
| 49 | exp dyspepsia/ or exp peptic ulcer/ or exp stomach ulcer/ | exp dyspepsia/ or exp peptic ulcer/ or exp stomach ulcer/ | exp Dyspepsia/ or exp Gastrointestinal Ulcers/ | (MH "Peptic Ulcer+" ) or (MH "Dyspepsia") | - |
| 50 | (dyspep* or (acid adj5 reflux)).ti,ab. | (dyspep* or (acid adj5 reflux)).ti,ab. | (dyspep* or (acid adj5 reflux)).ti,ab. | TI (dyspep* or (acid n4 reflux)) or AB (dyspep* or (acid n4 reflux)) | (TI=((dyspep* or (acid NEAR/4 reflux)) ) ) OR AB=((dyspep* or (acid NEAR/4 reflux)) ) |
| 51 | ((pep* adj5 ulcer*) or (stomach adj5 ulcer*) or (gastr* adj5 ulcer*).ti,ab. | ((pep* adj5 ulcer*) or (stomach adj5 ulcer*) or (gastr* adj5 ulcer*).ti,ab. | ((pep* adj5 ulcer*) or (stomach adj5 ulcer*) or (gastr* adj5 ulcer*).ti,ab. | TI ((pep* n4 ulcer*) or (stomach n4 ulcer*) or (gastr* n4 ulcer*)) or AB ((pep* n4 ulcer*) or (stomach n4 ulcer*) or (gastr* n4 ulcer*)) | (TI=((((pep* NEAR/4 ulcer*) or (stomach NEAR/4 ulcer*) or (gastr* NEAR/4 ulcer*)) ) ) OR AB=((((pep* NEAR/4 ulcer*) or (stomach NEAR/4 ulcer*) or (gastr* NEAR/4 ulcer*)) ) ) |
| 52 | exp Irritable Bowel Syndrome/ | exp Irritable Bowel Syndrome/ | exp Irritable Bowel Syndrome/ | MH "irritable bowel syndrome" | (TI=("irritable bowel syndrome")) OR AB=("irritable bowel syndrome") |
| 53 | ((irritable or functional or spastic) adj1 (bowel or colon)).ti,ab. | ((irritable or functional or spastic) adj1 (bowel or colon)).ti,ab. | ((irritable or functional or spastic) adj1 (bowel or colon)).ti,ab. | TI ( (irritable or functional or spastic) n1 (bowel or colon)) ) or AB ( (irritable or functional or spastic) n1 (bowel or colon)) ) | (TI=((((irritable or functional or spastic) NEAR/1 (bowel or colon)) ) ) OR AB=((((irritable or functional or spastic) NEAR/1 (bowel or colon)) ) ) |
| 54 | ibs.ti,ab. | ibs.ti,ab. | ibs.ti,ab. | TI ibs or AB ibs | (TI=(ibs)) OR AB=(ibs) |
| 55 | exp Fibromyalgia/ | exp Fibromyalgia/ | exp Fibromyalgia/ | MH fibromyalgia | - |
| 56 | fibromyalgia*.ti,ab. | fibromyalgia*.ti,ab. | fibromyalgia*.ti,ab. | TI fibromyalgia* or AB fibromyalgia* | (TI=(fibromyalgia*)) OR AB=(fibromyalgia*) |
| 57 | Enthesopathy/ | Enthesopathy/ | exp joint disorders/ | MH enthesopathy | (TI=(enthesopathy)) OR AB=(enthesopathy) |
| 58 | ((musculoskeletal or joint*) adj3 pain*).ti,ab. | ((musculoskeletal or joint*) adj3 pain*).ti,ab. | ((musculoskeletal or joint*) adj3 pain*).ti,ab. | TI ((musculoskeletal or joint*) n2 pain*) or AB ((musculoskeletal or joint*) n2 pain*) | TI=((((musculoskeletal or joint*) NEAR/2 pain*)) OR AB=((musculoskeletal or joint*) NEAR/2 |
|  |  |  |  |  | pain*)) |
| 59 | exp musculoskeletal pain/ | exp musculoskeletal pain/ | exp musculoskeletal disorders/ | (MH "Musculoskeletal Pain") or (MH "Back Pain") | - |
| 60 | exp Dermatitis, Atopic/ | exp atopic dermatitis/ | exp Dermatitis/ | (MH "Dermatitis, Atopic") | - |
| 61 | exp eczema/ | exp eczema/ | exp Eczema/ | (MH "Eczema") | - |
| 62 | (atopic adj3 dermatit*).ti,ab. | (atopic adj3 dermatit*).ti,ab. | (atopic adj3 dermatit*).ti,ab. | TI (atopic n2 dermatit*) or AB (atopic n2 dermatit*) | (TI=((atopic NEAR/2 dermatit*))) OR AB=((atopic NEAR/2 dermatit*)) |
| 63 | eczema.ti,ab. | eczema.ti,ab. | eczema.ti,ab. | TI (eczema) or AB (eczema) | TI=(eczema) OR AB=(eczema) |
| 64 | exp acne vulgaris/ or acne.ti,ab. | exp acne vulgaris/ or acne.ti,ab. | acne.ti,ab. | (MH "acne vulgaris") or TI (acne) or AB (acne) | TI=(acne) OR AB=(acne) |
| **Search Category 3. Longitudinal Studies** |  |  |  |  |  |
| 65 | exp cohort studies/ or cohort\$.tw. | exp cohort analysis/ or cohort\$.tw. | exp Cohort Analysis/ or cohort\$.tw. | TI ((cohort n3 stud*) or (cohort n1 analysis)) or AB ((cohort n3 stud*) or (cohort n1 analysis)) | ((TI=((cohort NEAR/3 stud*) or (cohort NEAR/1 analysis)))) OR AB=((cohort NEAR/3 stud*) or (cohort NEAR/1 analysis)))) |
|  | (longitudinal adj3 stud*).ti,ab. | exp longitudinal study/ or (longitudinal adj3 stud*).ti,ab. | exp Longitudinal Studies/ or (longitudinal adj3 stud*).ti,ab. | TI ((longitudinal n3 stud*) or (longitudinal n1 research)) or AB ((longitudinal n3 stud*) or (longitudinal n1 research)) | TI=((longitudinal NEAR/3 stud*) or (longitudinal n1 research)) ) ) OR AB=((longitudinal NEAR/3 stud*) or (longitudinal n1 research)) ) |
| 66 | (prospective adj3 stud*).ti,ab. | exp prospective study/ or (prospective adj3 stud*).ti,ab. | exp Prospective Studies/ or (prospective adj3 stud*).ti,ab. | (MH "Prospective Studies+") | TI=( prospective NEAR/3 stud*)) OR AB=(prospective NEAR/3 stud*)) |
| 68 | (follow-up adj3 stud*).ti,ab. | exp follow up/ or (follow-up adj3 stud*).ti,ab | (follow-up adj3 stud*).ti,ab. | TI (("follow up" n2 stud*) or (follow-up n2 stud*)) or AB (("follow up" n2 stud*) or (follow-up n2 stud*)) | TI=((("follow up" NEAR/2 stud*) or (follow-up NEAR/2 stud*)) ) ) OR AB=((("follow up" NEAR/2 stud*) or (follow-up NEAR/2 stud*)) ) |
| 69 | epidemiologic methods/ or controlled clinical trial.pt | - | - | (MH "Nonexperimental Studies+") OR (MH "Quasi-Experimental Studies+") |  |

